# The power of TOPMed imputation for the discovery of Latino enriched rare variants associated with type 2 diabetes

**DOI:** 10.1101/2022.09.30.22280535

**Authors:** Alicia Huerta-Chagoya, Philip Schroeder, Ravi Mandla, Aaron J. Deutsch, Wanying Zhu, Lauren Petty, Xiaoyan Yi, Joanne B. Cole, Miriam S. Udler, Peter Dornbos, Bianca Porneala, Daniel DiCorpo, Ching-Ti Liu, Josephine H. Li, Lukasz Szczerbiński, Varinderpal Kaur, Joohyun Kim, Yingchang Lu, Alicia Martin, Decio L. Eizirik, Piero Marchetti, Lorella Marselli, Ling Chen, Shylaja Srinivasan, Jennifer Todd, Jason Flannick, Rose Gubitosi-Klug, Lynne Levitsky, Rachana Shah, Megan Kelsey, Brian Burke, Dana M. Dabelea, Jasmin Divers, Santica Marcovina, Lauren Stalbow, Ruth J.F. Loos, Burcu F. Darst, Charles Kooperberg, Laura M. Raffield, Christopher Haiman, Quan Sun, Joseph B. McCormick, Susan P. Fisher-Hoch, Maria L. Ordoñez, James Meigs, Leslie J. Baier, Clicerio González-Villalpando, Maria Elena González-Villalpando, Lorena Orozco, Andrés Moreno, Carlos A. Aguilar-Salinas, Teresa Tusié, Josée Dupuis, Maggie C.Y. Ng, Alisa Manning, Heather M. Highland, Miriam Cnop, Robert Hanson, Jennifer Below, Jose C. Florez, Aaron Leong, Josep M. Mercader

**Affiliations:** Programs in Metabolism and Medical & Population Genetics, Broad Institute of Harvard and MIT, Cambridge, USA; Departamento de Medicina Genómica y Toxicología Ambiental, Instituto de Investigaciones Biomédicas, Universidad Nacional Autónoma de México, Mexico, Mexico; Unidad de Biología Molecular y Medicina Genómica, Instituto Nacional de Ciencias Médicas y Nutrición, México, Mexico; Center for Genomic Medicine, Massachusetts General Hospital, Boston, USA; Diabetes Unit, Massachusetts General Hospital, Boston, USA; Department of Medicine, Massachusetts General Hospital, Boston, USA; Vanderbilt Genetics Institute, Vanderbilt University Medical Center, Nashville, 37232, USA; ULB Center for Diabetes Research, Medical Faculty, Université Libre de Bruxelles, Brussels, 1070, Belgium; Department of Medicine, Harvard Medical School, Boston, USA; Division of Endocrinology, Boston Children’s Hospital, Boston, USA; Division of General Internal Medicine, Massachusetts General Hospital, Boston, USA; Department of Biostatistics, Boston University School of Public Health, Boston, USA; Department of Endocrinology, Diabetology and Internal Medicine, Medical University of Bialystok, Bialystok, 15-276, Poland; Clinical Research Centre, Medical University of Bialystok, Bialystok, 15-276, Poland; Vanderbilt Genetics Institute, Division of Genetic Medicine, Vanderbilt University Medical Center, Nashville, 37232, USA; Analytic and Translational Genetics Unit, Massachusetts General Hospital, Boston, USA; WELBIO, Université Libre de Bruxelles, 1070, Belgium; Department of Clinical and Experimental Medicine, and AOUP Cisanello University Hospital, University of Pisa, Pisa, 56126, Italy; Department of Pediatrics, University of California San Francisco, San Francisco, USA; Department of Pediatrics, University of Vermont, Burlington, USA; Department of Pediatrics, Boston Children’s Hospital, Boston, USA; Pediatric Endocrinology, Diabetes, and Metabolism, Case Western Reserve University and Rainbow Babies and Children’s Hospital,, Cleveland, USA; Department of Pediatrics, Division of Pediatric Endocrinology and Pediatric Diabetes Center, Massachusetts General Hospital, Boston, USA; Pediatric Endocrinology and Diabetes, Children’s Hospital of Philadelphia, Philadelphia, USA; Pediatric Endocrinology, University of Colorado School of Medicine, Aurora, USA; Biostatistics Center, The George Washington University, Rockville, USA; Department of Epidemiology, University of Colorado School of Medicine, Aurora, USA; NYU Langone Health, New York, USA; Medpace Reference Laboratories, Cincinnati, USA; The Charles Bronfman Institute of Personalized Medicine, Icahn School of Medicine at Mount Sinai, New York, USA; Novo Nordisk Foundation Center for Basic Metabolic Research, Faculty of Health and Medical Science, University of Copenhagen, Copenhagen, USA; Division of Public Health Science, Fred Hutchinson Cancer Center, Seattle, USA; Department of Genetics, University of North Carolina at Chapel Hill, Chapel Hill, USA; Department of Population and Public Health Sciences, University of Southern California, Los Angeles, USA; Norris Comprehensive Cancer Center, University of Southern California, Los Angeles, USA; Department of Biostatistics, University of North Carolina at Chapel Hill, Chapel Hill, USA; School of Public Health, The University of Texas Health Science Center at Houston, Brownsville, 78520, USA; Unidad de Biología Molecular y Medicina Genómica, Instituto Nacional de Ciencias Médicas y Nutrición, Mexico, Mexico; Phoenix Epidemiology and Clinical Research Branch, National Institute of Diabetes and Digestive and Kidney Diseases, National Institutes of Health, Phoenix, USA; Centro de Estudios en Diabetes, Unidad de Investigacion en Diabetes y Riesgo Cardiovascular, Centro de Investigacion en Salud Poblacional, Instituto Nacional de Salud Pública, Mexico, Mexico; Laboratorio Inmunogénomica y Enfermedades Metabólicas, Instituto Nacional de Medicina Genómica, Mexico, Mexico; Laboratorio Nacional de Genómica para la Biodiversidad, Unidad de Genómica Avanzada, Irapuato, Mexico; Unidad de Investigación de Enfermedades Metabólicas y Dirección de Nutrición, Instituto Nacional de Ciencias Médicas y Nutrición Salvador Zubirán, Mexico, Mexico; Clinical and Translational Epidemiology Unit, Massachusetts General Hospital, Boston, USA; Department of Epidemiology, University of North Carolina at Chapel Hill, Chapel Hill, USA; ULB Center for Diabetes Research, Medical Faculty, Université Libre de Bruxelles, 1070, Belgium; Division of Endocrinology, Erasmus Hospital, Université Libre de Bruxelles, 1070, Belgium; Diabetes Epidemiology and Clinical Research Section, National Institute of Diabetes and Digestive and Kidney Diseases, Phoenix, USA; Endocrine Division, Massachusetts General Hospital, Boston, USA

**Author notes:** **Corresponding author:** Josep M Mercader, Programs in Metabolism and Medical and Population Genetics, Broad Institute of Harvard and MIT, 75 Ames St, 02142, Cambridge, MA, United States of America. These authors jointly directed this work (Jose C. Florez, Aaron Leong, Josep M. Mercader).

## Abstract

**Hypothesis:** The prevalence of type 2 diabetes is higher in Latino populations compared with other major ancestry groups. Not only has the Latino population been systematically underrepresented in large-scale genetic analyses, but previous studies relied on the imputation of ungenotyped variants based on the 1000 Genomes (1000G) imputation reference panel, which results in suboptimal capture of low-frequency or Latino-enriched variants. The NHLBI Trans-Omics for Precision Medicine (TOPMed) reference panel represents a unique opportunity to analyze rare genetic variations in the Latino population.

**Methods:** We evaluate the TOPMed imputation performance using genotyping array and whole-exome sequence data in 6 Latino cohorts. To evaluate the ability of TOPMed imputation of increasing the identified loci, we performed a Latino type 2 diabetes GWAS meta-analysis in 8,150 type 2 diabetes cases and 10,735 controls and replicated the results in 6 additional cohorts including whole-genome sequence data from the *All of Us* cohort.

**Results:** We show that, compared to imputation with 1000G, the TOPMed panel improves the identification of rare and low-frequency variants. We identified 26 distinct signals including a novel genome-wide significant variant (minor allele frequency 1.6%, OR=2.0, P=3.4×10^−9^) near *ORC5*. A Latino-tailored polygenic score constructed from our data and GWAS data from East Asian and European populations improves the prediction accuracy in a Latino target dataset, explaining up to 7.6% of the type 2 diabetes risk variance.

**Conclusions:** Our results demonstrate the utility of TOPMed imputation for identifying low-frequency variation in understudied populations, leading to the discovery of novel disease associations and the improvement of polygenic scores.

## INTRODUCTION

Latino is a diverse ethnic group recently admixed from Native American, European, and African ancestries with a high prevalence of metabolic disorders, including type 2 diabetes. Although genetic studies in the Latino populations are limited, they have revealed unexpected pathways and potential therapeutic targets for type 2 diabetes.[1–4] This is the case for a Native American haplotype within the *SLC16A11* gene, which was identified as the main genetic contributor to type 2 diabetes in Latino people[1, 4], a rare risk variant within *HNF1A* unique to Latino populations[2] and a loss-of-function (LoF) Latino-enriched variant within *IGF2* associated with a 20% reduced risk of type 2 diabetes.[3]

Unlike genetically homogenous populations, the complex linkage disequilibrium (LD) structure of admixed populations imposes challenges in implementing statistical methods that are crucial to maximize genetic discoveries.[5] This is especially relevant for genotype imputation, a method to estimate the genotype probabilities at genetic variants that have not been experimentally genotyped.[6] A major factor limiting the accuracy of genotype imputation in Latino samples has been the poor representation of their haplotypes in available reference panels. Until recently, the only available panel for the imputation of Latino samples was the 1000G reference panel, whose latest version includes only 352 Latino samples out of 2,504 total samples.[7] The multi-ancestry NHLBI Trans-Omics for Precision Medicine (TOPMed) program has released a reference panel for genotype imputation which includes the highest sequencing coverage (*i*.*e*. 30x) and the largest number of reference samples (*i*.*e*. 97,256) to date, of whom ∼15% are Latino individuals. It has shown to increase the number of well-imputed low-frequency variants in the admixed Latino cohort HCHS/SOL.[8, 9]

We hypothesized that by boosting the identification of variants in Latino samples with the recently released TOPMed reference panel, we would consequently improve our knowledge of the genetic architecture of type 2 diabetes in the Latino population. We show that TOPMed improves the coverage of genetic variation, especially within the rare and low-frequency spectrum, in 6 Latino cohorts. We then performed a type 2 diabetes GWAS meta-analysis in this population and identified previously unreported genetic signals. We also performed association analyses on a collection of type 2 diabetes-related phenotypes from TOPMed Latino imputed datasets to allow the interpretation of our novel genetic variants that have low frequencies or are absent in other publicly available biobanks, which mainly contain individuals of European ancestry. Finally, we leveraged the generated GWAS data to develop, in combination with GWAS datasets from other ancestries, an improved type 2 diabetes polygenic score (PS) for the Latino population.

## SUBJECTS AND METHODS

### Discovery sample

A detailed description of the methods is found in the electronic supplementary material. We aggregated data from 6 Latino cohorts with a total sample size of 18,885 individuals (8,150 cases and 10,735 controls): the Slim Initiative for Genomic Medicine in the Americas (SIGMA) Cohorts[1–3], the Mexican Biobank Cohort (MXBB)[10], the Mass General Brigham (MGB) Biobank[11] and the Genetic Epidemiology Research on Aging Cohort (GERA)[12] (Figure 1, Table S1).

**Fig. 1.**
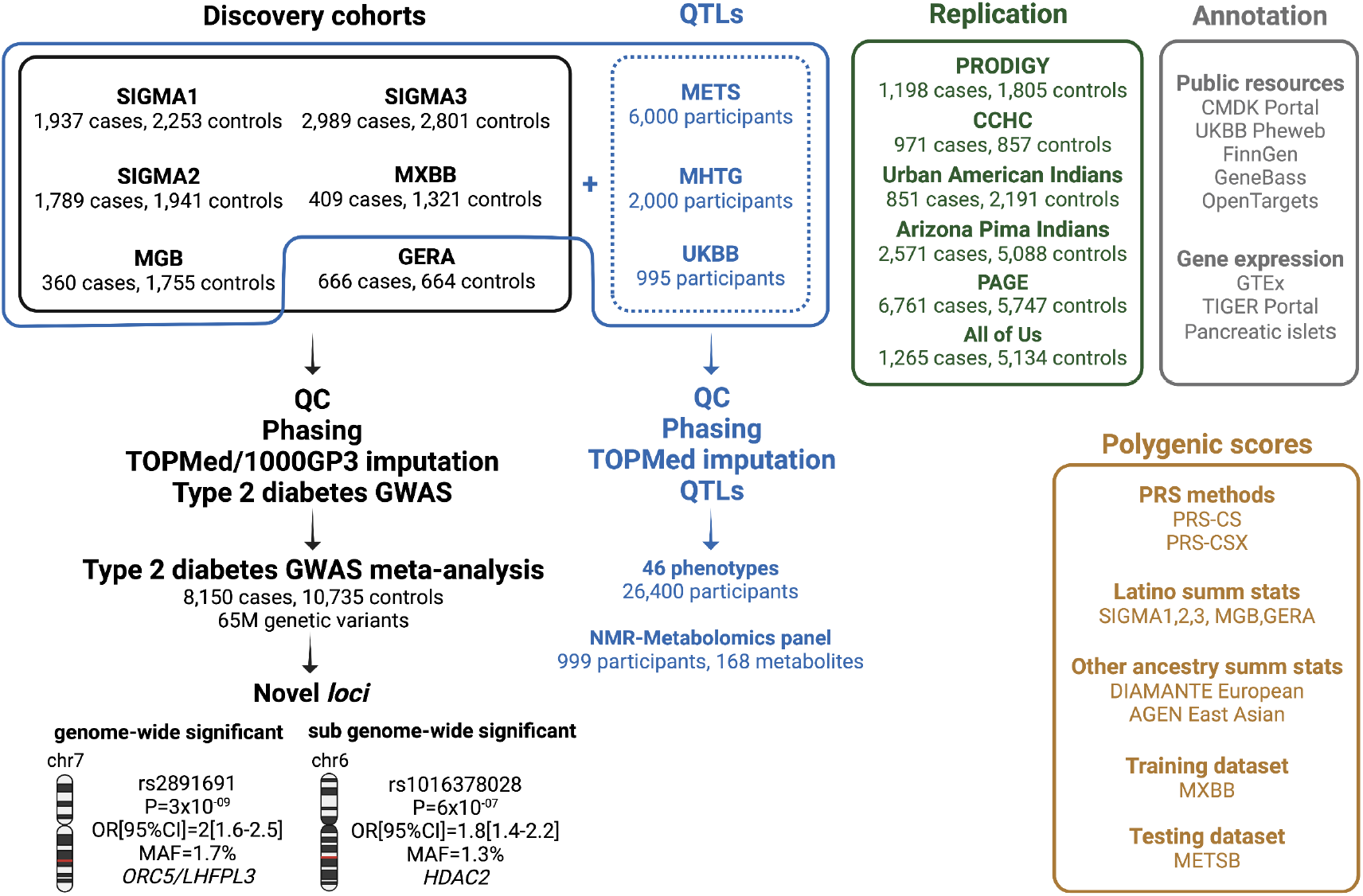
General overview of the study. Six cohorts of admixed Latino ancestry, representing a total of 8,150 type 2 diabetes cases and 10,735 controls, were imputed with the TOPMed and 1000G Phase 3 panels (black box). A type 2 diabetes GWAS meta-analysis of the imputed variants resulted in the identification of two novel loci, which were tested for replication in six additional Latino cohorts (green box). They were also interrogated for association with a collection of phenotypes in eight Latino cohorts (blue box) and for functional evidence in multiple available resources (grey box). The generated Latino type 2 diabetes GWAS data was used, in combination with GWAS from other ancestries, to construct ancestry-specific and cross-population type 2 diabetes polygenic scores (brown box).

We selected Latino samples based on their genetically estimated ancestry and calculated the genetic-based global ancestry using Admixture[13] assuming five ancestral populations (k=5) (Figure S1). All human research was approved by the relevant Institutional Review Boards and conducted according to the Declaration of Helsinki. All participants provided written informed consent.

### Genotyping, quality control and imputation

Genotyping was done using several commercially available genome-wide arrays, and for a subset of the samples (N=9,520), we integrated whole-exome sequencing (WES) (Table S1). We applied pre-imputation quality control to each dataset separately. Clean datasets were phased using SHAPEIT2[14] and used as input for imputation. For comparison purposes, we imputed the phased haplotypes using both 1000G Phase 3 version 5[15] and TOPMed reference panels freeze 8[8].

### Imputation performance evaluation

We evaluated the performance of TOPMed and 1000G imputations by summarizing the chromosome-wise r^2^ quality measure and the number of well-imputed (r^2^≥0.8) variants at different allele frequency thresholds. We used available WES data from SIGMA3 cohort and estimated the proportion of the sequenced variants, for chromosome 22 only, that were well-imputed with TOPMed and 1000G panels at different WES allele frequency thresholds. We used SnpEff[16] to annotate the WES variants and estimated the percentage of well-imputed variants identified with TOPMed and 1000G imputations. We also calculated the effective sample size needed to reach 80% statistical power to detect genome-wide significant associated signals (∝=5×10^−8^) at different effect sizes and allele frequencies covered by the imputations.

### Type 2 diabetes association and meta-analysis

Association analyses were performed in each cohort with SNPTEST[17]. Models were adjusted for sex, age, BMI and 10 PCs to account for population structure. We ran additional models without adjusting for BMI. Only well-imputed variants (r^2^≥0.5) were meta-analyzed using the inverse of the corresponding squared standard errors in METAL.[18] The statistical significance threshold was set to *P*<5×10^−8^.

To extract the distinct type 2 diabetes associated signals, we clumped all variants with an association *P*-value<5×10^−6^. We set an LD r^2^=0.5 and a distance between variants of 250 kb. If the variant was located within a previously reported type 2 diabetes-related *locus*, we used a conditioning strategy to test for distinct signals. Distinct variants with sub-genome-wide significance (*P*<1×10^−6^) that were only imputed with the TOPMed reference panel, showed increased frequency in the Latino population and were > 250 kb from other reported genome-wide significant variants from large consortia analyzing either European or East Asian populations[19, 20] were considered for further investigation.

### Replication sample

Variants associated with type 2 diabetes risk at genome-wide and sub genome-wide significance were tested for replication in six independent cohorts: the Cameron County Hispanic Cohort (CCHC)[21], the Urban American Indians and Arizona Pima Indians cohorts[22], the Population Architecture using Genomics and Epidemiology (PAGE) study[23], the All of Us Research Program[24] and the Progress in Diabetes Genetics in Youth (PRODIGY), which comprises the Treatment Options for Type 2 Diabetes in Adolescents and Youth (TODAY)[25], the SEARCH for Diabetes in Youth studies[26], the Type 2 Diabetes Genetics Exploration by Next-generation sequencing in multi-Ethnic Samples (T2D-GENES) cohorts and the Mexican Metabolic Syndrome (METS) Cohort.[27] (Table S2).

### Quantitative Trait Locus (QTL) analyses

Given the lack of large-scale publicly available biobanks with Latino samples that may allow for better characterization of our novel signals, we assembled a collection of cohorts to perform QTL analyses focused on 46 glycemic, anthropometric and lipid traits. In addition to 5 of the Latino cohorts analyzed in the type 2 diabetes meta-analysis (*i*.*e*. SIGMA1, SIGMA2, SIGMA3, MXBB and MGB Biobank), we included three extra cohorts, which we also imputed to the TOPMed panel: the METS Cohort, the Mexican Hypertriglyceridemia (MHTG) Cohort, as well as the genetically identified Latino samples from the UK Biobank (UKBB).[28] We also analyzed the Nightingale Nuclear Magnetic Resonance-based panel of 168 metabolomic biomarkers in Latino samples from the UKBB. Association analyses were done with a maximum of 26,400 adult Latino individuals depending on the trait, of whom 19,459 were diabetes-free.

### Credible sets

For each novel variant, we identified the set of variants with 99% probability of containing the causal variant. We used a Bayesian method[29], considering variants in LD with the lead variant (r^2^>0.1). We calculated LD using genetic data from 1,996 Hispanic/Latino samples from TOPMed Freeze 5b.

### Genomic annotation

We used the 99% credible sets for each novel signal to annotate their genomic effect using the VEP[30] (GRCh38.p7) and SNPNEXUS[31] applications. We used GTEx V8[32] to assess the influence of the variants in gene-level expression, as well as the TIGER Portal[33] for evaluating the gene-level expression in pancreatic islets and the Islet Gene View[34] for assessing the gene co-expression in human islets. We also assessed individual variant associations with a variety of common phenotypes and diseases using the Common Metabolic Disease Knowledge Portal (cmdgenkp.org. 2021 Nov 15), as well as other resources.

### Expression of genes near novel variants

We assessed the expression levels of the genes ± 500 kb around the novel signals in human islets under different conditions pertaining to type 1 diabetes and type 2 diabetes. Gene expression differences between groups were assessed using *P*-values and adjusted *P*-values (Benjamini Hochberg correction) determined by the Wald test using the DESeq2 pipeline.[35] Transcript per million (TPM) was normalized by Salmon 1.4.0.[36]

### Polygenic scores

Polygenic scoring using single ancestry summary statistics and LD reference panels was calculated via Bayesian Regression and Continuous Shrinkage priors as implemented in PRS-CS.[37] We used the UKBB LD reference panel and GWAS summary statistics from European[19], East Asian[20] and Latin American populations. GWAS Latino summary statistics were calculated using a meta-analysis with five of the discovery cohorts (*i*.*e*. SIGMA1, SIGMA2, SIGMA3, MGB, and GERA). Then, we used the estimated posterior SNP effect sizes for each ancestry to calculate and evaluate the performance of the PSs in a training cohort (*i*.*e*. MXBB). The best model was tested in a target cohort (*i*.*e*. the METS Cohort).

Given that the ancestry-specific PSs were not highly correlated (r^2^<0.3), we also used PRS-CSx[38], a novel method that improves cross-population polygenic prediction by integrating GWAS summary statistics from multiple populations. We assessed the performance of the ancestry-specific versus the cross-population PS.

## RESULTS

### Overall analysis strategy

Figure 1 summarizes our overall analysis strategy. We meta-analyzed 6 type 2 diabetes GWAS of Latin American ancestry, comprising a total of 8,150 cases and 10,735 controls. Individuals were from hospital and population-based studies. All cohorts were imputed with TOPMed and 1000G Phase 3 panels and the imputation performance was evaluated. To replicate the novel loci, we analyzed 13,595 type 2 diabetes cases and 23,403 controls from 6 independent cohorts of Latino ancestry. To gain further insight into the novel loci, especially for variants enriched in Latin American ancestry, we created a Latin American collection of type 2 diabetes-related phenotypes that included a total of 26,400 Latino participants with 46 available glycemic and anthropometric traits, as well as 168 metabolomic traits. We used publicly available resources to interrogate our top signals, including functional annotation of the credible sets and evaluating gene expression of nearby genes in pancreatic islets from either type 1 diabetes or type 2 diabetes cases and controls or treated under conditions relevant for diabetes pathophysiology. We then used the generated Latino GWAS data, in combination with GWAS from other ancestries, to construct ancestry-specific or cross-population type 2 diabetes PSs (Figure 1).

### TOPMed imputation performance

Across all cohorts, imputation using the TOPMed reference panel resulted in 41M high quality (r^2^≥0.8) variants on average, of which 24M were rare (MAF<0.1%) representing a 6.5-fold increased number of rare variants imputed with TOPMed compared to 1000G (Figure 2a). For variants that were imputed by both panels in all cohorts, the quality of imputation consistently improved when using TOPMed, particularly for low-frequency and rare variants (Figure 2b).

**Fig. 2.**
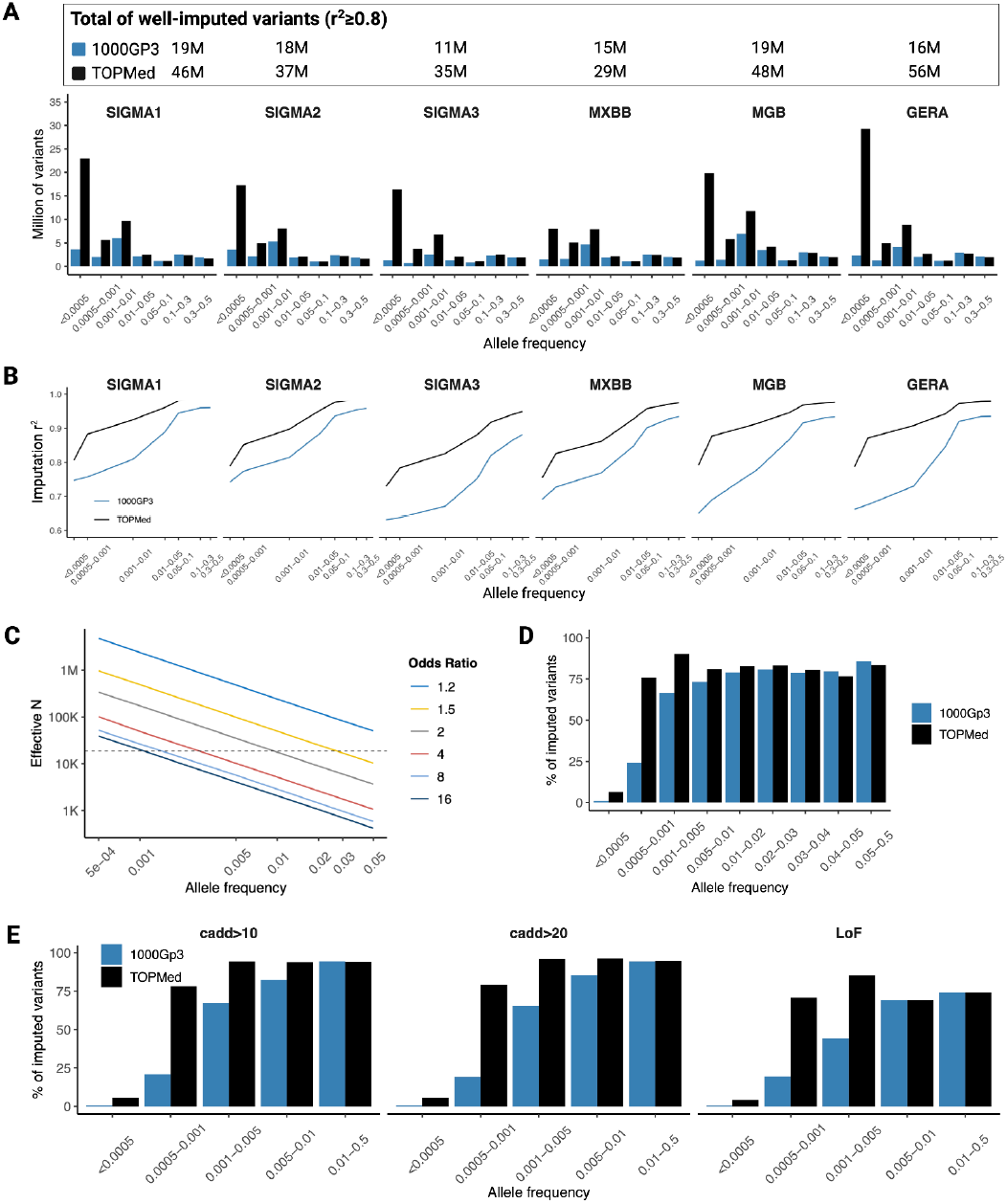
Performance of the TOPMed reference panel for the imputation of Latino samples. a. Number of chromosome-wide well-imputed variants (imputation r^2^≥0.8) by allele frequency for each analyzed cohort when using the 1000 Genomes Phase 3 (blue) or the TOPMed (black) reference panels. b. Average chromosome-wide imputation quality by allele frequency for each analyzed cohort when using the 1000 Genomes Phase 3 (blue) or the TOPMed (black) reference panels. c. Effective sample size required for reaching 80% statistical power to detect genome-wide significant signals at different effect sizes (OR). The dotted lines show the discovery effective sample size of this study (N=18,531) d. Percentage of the exome sequenced variants in chromosome 22 that could be imputed when using the 1000 Genomes Phase 3 (blue) or the TOPMed (black) reference panels. e. Percentage of the exome sequenced LoF and deleterious CADD-predicted variants in chromosome 22 that could be imputed when using the 1000 Genomes Phase 3 (blue) or the TOPMed (black) reference panels.

The improvement of TOPMed imputation to detect low-frequency and rare variation was confirmed using WES data. The TOPMed panel allowed the identification of >80% of the sequenced variants with a WES MAF≥0.1% compared to 60% for the same MAF cutoff with the 1000G panel (Figure 2d). The TOPMed panel also improved the identification of likely pathogenic variants predicted as deleterious, which usually occur at low frequency (Figure 2e).

### Type 2 diabetes GWAS meta-analysis

To illustrate the gain in discovery when using TOPMed imputation, we tested genetic variants for association with type 2 diabetes in 6 cohorts, meta-analyzing 65M variants with an imputation r^2^ quality ≥0.5. Our discovery sample comprised a total of 18,885 Latino non-related individuals (8,150 cases, 10,735 controls) (Figure 1, Table S1, Figure S1).

We identified 26 distinct variants at 13 loci associated with type 2 diabetes at a standard genome-wide significance threshold of *P*<5×10^−8^. Among them, we replicated 25 previously reported type 2 diabetes-associated common variants, including those consistently identified in multiple populations (*e*.*g*. variants at *KCNQ1* and *TCF7L2*) and others enriched in the Latino population (*e*.*g*. variants at *SLC16A11*) (Figure 3a, Table S3, Figure S2).

**Fig. 3.**
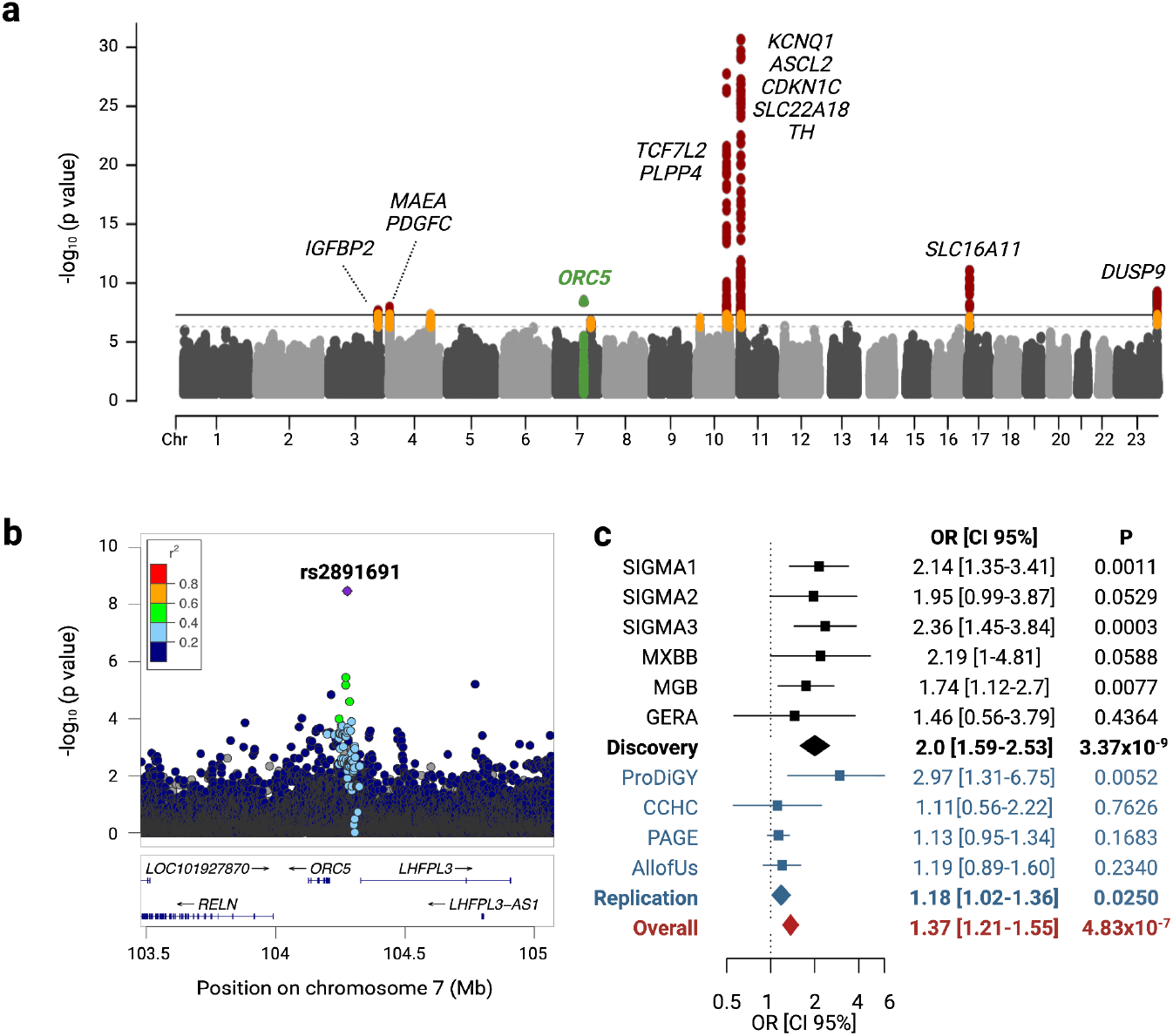
Type 2 diabetes GWAS meta-analysis in Latino population. a. Manhattan plot of the meta-analysis association statistics highlighting the loci with genome-wide significance (red) or sub genome-wide significance (orange) for type 2 diabetes. b. Regional association plot of the novel *ORC5/LHFPL3 locus* associated with type 2 diabetes risk. c. Forest plot of the GWAS association statistics for the novel *ORC5/LHFPL3 locus* in the discovery (black) and the replication (blue) cohorts.

We identified a novel *locus* between the *ORC5* and *LHFPL3* genes on chromosome 7. The intergenic index variant, rs2891691, has low frequency in Latino people and is associated with 2-fold increased odds of developing type 2 diabetes (MAF=1.7%, OR [95% CI] =2.0 [1.59-2.52], *P*=3.4×10^−9^) (Figure 3b,c).

Although it was also imputed with the 1000G panel, TOPMed’s higher imputation quality strengthened the association (1000G: mean±SD imputation r^2^=0.948 ± 0.057, *P*=2.3×10^−8^; TOPMed: mean ± SD imputation r^2^=0.983 ± 0.009, *P*=3.4×10^−9^). This variant is rare in Europeans, yet prevalent among African and East Asian populations. However, no association with type 2 diabetes has been reported in these populations. The lack of association in African ancestry may suggest that the lead variant we identified is in LD with the causal variant in the Latin American but not in African or East Asian populations, a phenomenon also observed in other multi-ancestry meta-analyses.[39] It can also be explained by lack of power, or differences in environmental exposures in these populations.

A sex-dimorphism in *RELN* gene expression has been documented, with higher *RELN* expression in women[40] and sex hormones likely mediating *RELN* expression. Because of the proximity of *RELN* to rs2891691, we evaluated the sex-specific association with type 2 diabetes and also tested for heterogeneity between sex-specific allelic effects using GWAMA.[41] We found that rs2891691 showed a larger effect and was more associated with type 2 diabetes in women (OR [95% CI] =2.4 [1.73-3.22], *P*=6.6×10^−8^) compared to men (OR [95% CI] =1.5 [1.08-2.19], *P*=0.018), yet the between-sex heterogeneity did not reach statistical significance (*P*=0.076) (Table S5).

### Replication analysis

The replication analysis comprised 13,617 type 2 diabetes cases and 20,822 controls (Table S2). We assessed the association of rs2891691 with type 2 diabetes in the four replication cohorts it was present. The meta-analysis of all replication cohorts was nominally significant and showed a consistent direction of effect with the discovery sample (OR [95% CI] =1.18 [1.02-1.36], *P*=0.025) (Figure 3c, Table S4).

By querying our Latin American collection of type 2 diabetes-related phenotypes we found that the rs2891691 type 2 diabetes risk allele C was nominally associated with lower fasting glucose levels (beta [95% CI] =-0.18 [-0.02 - -0.034] mg/dl, *P*=0.026) (Table S7).

The 99% credible set consisted only of the index variant rs2891691 (Table S9), which is in a quiescent or repressed chromatin state in diabetes-relevant tissues, such as islets of Langerhans, adipocytes, skeletal muscle, and adipose tissue (ChromHMM 13-state model). However, this variant (chr7:104276872:C:A, hg38) is located in a chromatin region (chr7:104276536-104277258) that shows stable open chromatin status after 2 or 4h of IFNα exposure in EndoC-βH1 cells, a human beta cell line.[42, 43] IFNα treatment induces endoplasmic reticulum stress in human islets and EndoC-βH1 cells, consequently reducing the insulin content with a rise in the proinsulin:insulin ratio.[44]

To better characterize the role of the novel *locus ORC5*/*LHFPL3*, we assessed gene expression using the GTEx[32] and TIGER[33] Portals. We found that the *ORC5* gene is expressed ubiquitously, while *LHFPL3* is specifically expressed in the brain (Figure S4a,b). We then assessed the expression levels of genes ± 500 kb around the novel signal in human islets under different conditions relevant to diabetes pathophysiology. *ORC5* was expressed in human islets under basal conditions (Figure S5a). It was downregulated after 2h and 8h exposure to IFNα, and upregulated by exposure to brefeldin A, an endoplasmic reticulum and Golgi stress inducer that inhibits glucose-stimulated insulin secretion[45] (Figure S5c).

### Prioritizing sub-genome-wide significant variants with additional evidence

We next searched for variants whose association with type 2 diabetes was at sub-genome-wide significance (*P*<5×10^−6^) but that deserved further replication as they were enriched or unique of Latino population, and/or exclusively imputed with the TOPMed panel. (Figure 4a). We first identified 23 sub genome-wide distinct variants. Of these, three located in or near *TACC2, FGFR2*, and *CCND2* were in known type 2 diabetes loci but retained locus-wide significance (P<5×10^−5^) after conditioning on the nearest known associated variant. We consider these three variants as distinct from known type 2 diabetes-associated loci. In addition, three sub-genome-wide significant variants are located more than 1 Mb away from any previously reported type 2 diabetes association and are potentially novel (Figure 4a, Table S6). We prioritized rs1016378028, a low frequency variant associated with 1.77-fold increased risk of developing type 2 diabetes (MAF=1.3%, OR [95% CI] =1.77 [1.41-2.21], *P*=7.0×10^−7^), because it is a Latino private variant, only imputed with the TOPMed reference panel, lies in an intron of *HDAC2*, a gene under strong purifying selection (probability of being LoF intolerant [pLI]=1, gnomAD, accessed March 2022) and it is highly and specifically expressed in pancreatic islets (tiger.bsc.es, accessed March 2022).[33]

**Fig. 4.**
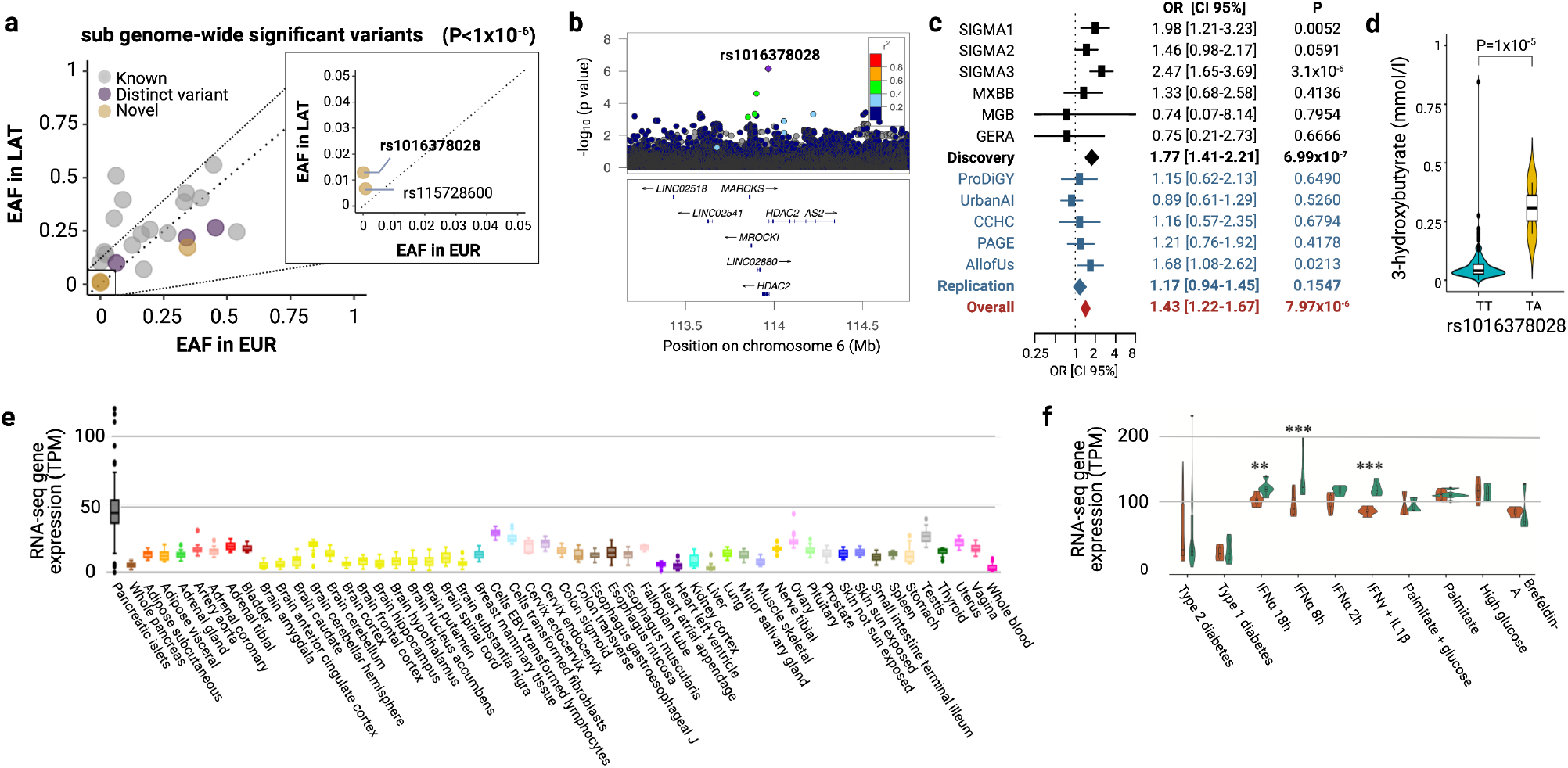
Sub genome-wide significant *HDAC2* novel type 2 diabetes loci. a. Scatter plot of the effect allele frequencies from the sub genome-wide significant variants in Latino *versus* European populations, highlighting those that are distinct from the known lead type 2 diabetes -associated variants (purple) and those that are in novel loci (yellow). b. Regional association plot of the novel *HDAC2 locus* associated with type 2 diabetes risk. c. Forest plot of the association statistics in the discovery (black) and the replication (blue) cohorts. d. Serum 3-hydroxybutyrate levels in non-carriers and carriers of rs1016378028 variant. *e. HDAC2* gene expression in multiple tissues from GTEx and TIGER portals. *f. HDAC2* gene expression in human islets from type 1 and type 2 diabetes and control islets treated or not with different cytokines or other stressor compounds. INF-α: interferon alpha; INF-γ: interferon gamma; IL1β: interleukin 1 beta. The presence of each condition is labeled in orange and the control group in green. Asterisks represent the adjusted *P*-values (Benjamini Hochberg correction). ** adjusted *P*<0.01, *** adjusted *P*<0.001.

Although the replication results did not show statistical significance, the direction of the effect was consistent with the discovery effect (OR [95% CI] =1.17 [0.94-1.45], *P*=0.1547) (Figure 4b,c, Table S4, S6). The variant is almost absent in non-Latino populations (MAF<0.01%). In the DIAMANTE European meta-analysis[46], a suggestive signal ∼80 kb upstream of rs1016378028 (rs4945979, *P*=4.8×10^−6^) was reported. After conditioning for the rs4945979 variant, statistical significance of our identified variant remained essentially the same (OR [95% CI] =1.75 [1.4-2.2], *P*=4.5×10^−7^).

By analyzing association with metabolites, we observed that the rs1016378028 risk allele was significantly associated with higher levels of acetone (beta [95% CI] =3.3[2.06-4.48], *P*=1.2×10^−7^), 3-hydroxybutyrate (beta [95% CI] =2.7[1.5-3.9], *P*=1.01×10^−5^) and acetoacetate (beta [95% CI] =2.5[1.3-3.6], *P*=3.3×10^−5^) (Figure 4d, Table S8). The risk allele was also nominally associated with lower hip circumference (beta [95% CI] =-0.1[-0.01 - -0.18], *P*=0.02) and higher waist-to-hip ratio (beta [95% CI] =0.13[0.01-0.24], *P*=0.03) (Table S7).

Because *HDAC2* is highly and specifically expressed in pancreatic islets, compared to other human tissues (GTEx[32] and TIGER[33] Portals) (Figure 4e), we examined the *HDAC2* gene expression in human islets exposed to different conditions and diseases states (*i*.*e*., type 1 diabetes and type 2 diabetes). Exposure of human islets to IFNα (8h log2-fold change=-0.38, *P*=6×10^−7^, 18h log2-fold change=-0.28, *P*=3×10^−4^) or IFNγ+IFNβ (log2-fold change=-0.39, *P*=3×10^−7^) showed downregulated *HDAC2* expression (Figure 4f). These cytokines mimic the proinflammatory milieu of type 1 diabetes, inhibit β cell function[47, 48], induce β cell stress and may trigger β cell dedifferentiation in type 2 diabetes.[49, 50]

### Development of PSs for the Latino population

We then developed a PS for type 2 diabetes in Latino people using our TOPMed imputed GWAS meta-analysis data. This PS explained 1.6% of type 2 diabetes status variance (Figure 5a), which is expected given the relatively small sample size of the GWAS used to construct the PS compared to European and East Asian summary statistics. The PS derived from the DIAMANTE European GWAS[46] and from AGEN East Asian GWAS[20] explained 5.1% and 4.4% of the type 2 diabetes variance in the Latino population, respectively. We observed that PSs of European, East Asian, and our Latino TOPMed meta-analysis showed a weak correlation (r^2^<0.2), suggesting that the three PSs could provide orthogonal information and improve the overall predictive performance. To develop a PS that incorporates GWAS data from the three ancestries, we used PRS-CSx[51], a method that allows for the integration of summary statistics and LD reference panels from different ancestries. The trans-ancestry PRS-CSx including our Latino GWAS, European, and East Asian GWAS summary statistics, explained 7.6% of the type 2 diabetes variance in the Latino target sample. Our Latino GWAS added 1% of the explained variance compared to the PS using only European and East Asian GWAS summary statistics, which explained 6.6% of the variance.

**Fig. 5.**
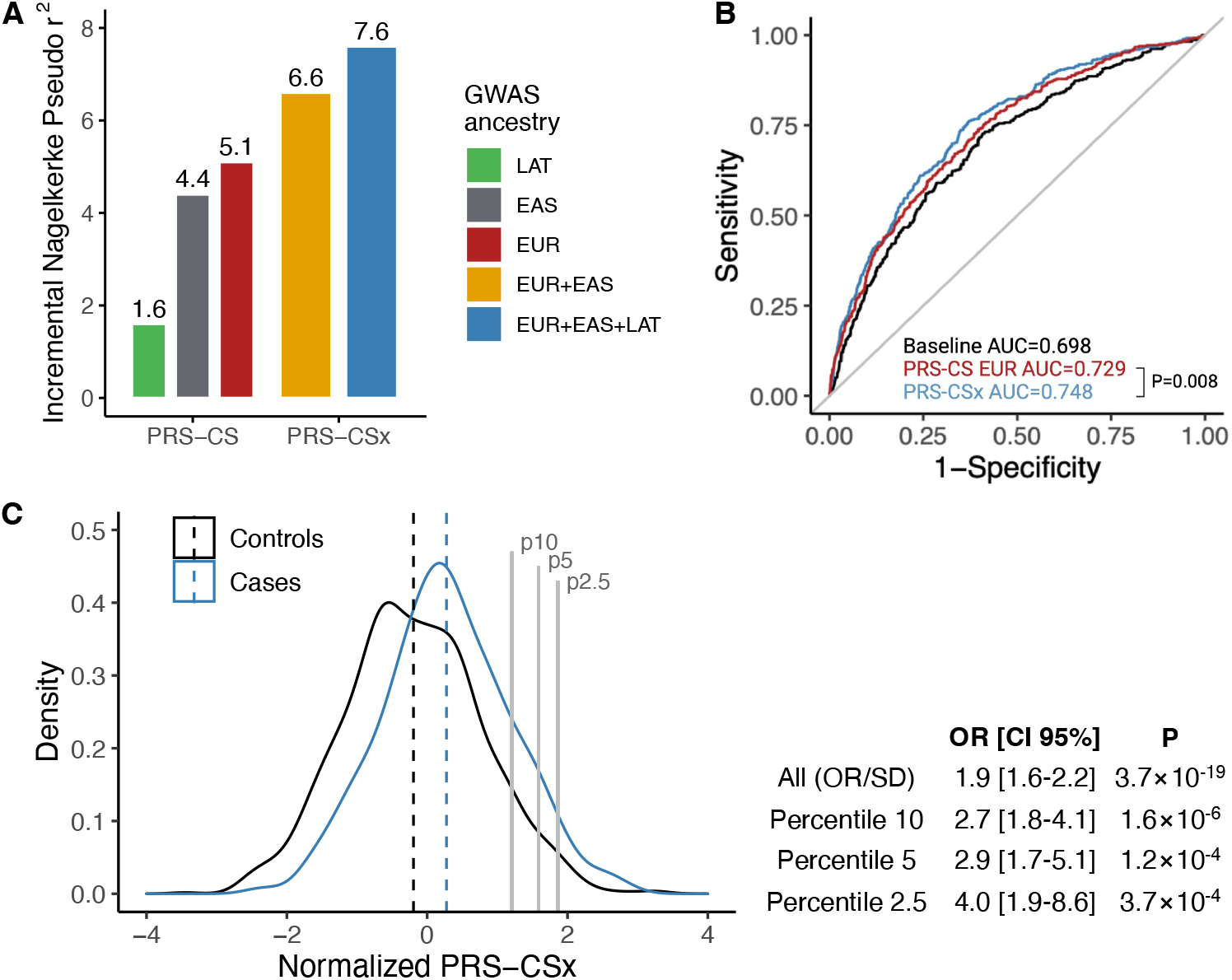
Polygenic Risk Score for the risk of type 2 diabetes in Latino population. a. Variance explained by a PS using these Latino GWAS association statistics (green), the AGEN East-Asian GWAS association statistics (red), the DIAMANTE European GWAS association statistics (grey), a combination of DIAMANTE European and AGEN East-Asian GWAS association statistics (yellow) and a combination of DIAMANTE European, AGEN East Asian and these Latino GWAS association statistics (blue). METSB was used as testing cohort. b. ROC curves for the type 2 diabetes risk prediction explained by a model including sex, age and 10 PCs of ancestry (black) and a model including the same covariates and a PS constructed using combination of DIAMANTE European, AGEN East Asian and these Latino GWAS association statistics (blue). c. Distribution of a cross-population PS using a combination of DIAMANTE European, AGEN East Asian and these Latino GWAS association statistics in type 2 diabetes cases (blue) and controls (black). Table shows the OR per standard deviation attributed to the cross-population PS, as well as the OR for high-risk individuals.

Each standard deviation of the trans-ancestry PRS-CSx was associated with an OR [95% CI] =1.9 [1.6-2.2], *P*=3.7×10^−19^. People in the 2.5 percentile of the PRS-CSx showed 4 times more risk of developing type 2 diabetes (OR [95% CI] =4.01 [1.87-8.62], *P*=3.7×10^−4^) (Figure 5c). The ROC AUC [95% CI] of the full model including the trans-ancestry PRS-CSx was 0.748 [0.72-0.775] compared to the AUC [95% CI] of 0.729 [0.701-0.758] of the PRS-CS including European GWAS summary statistics only, representing a 2% improvement in prediction accuracy (*P*=0.008).

## DISCUSSION

Latino populations have been systematically underrepresented in most genetic studies. Yet, recent studies of type 2 diabetes in Latino populations have been fruitful, even with sample sizes orders of magnitude smaller than those of European or East Asian ancestries. To date, the poor representation of Latino samples with genotype and phenotype data constrains nearly every step of a gene-disease association framework, including genotype imputation, a cost-effective technique to improve the resolution of a GWAS. This is more problematic for low-frequency and rare variation. Instead, next-generation sequencing technologies have typically been chosen, but are more expensive, precluding the study of large samples. This study was motivated by the recent release of the TOPMed imputation panel, which is the reference panel with the largest number of Latino haplotypes as compared to all available reference panels.

In this study, we aggregate genotype and WES data from 6 separate datasets to test the improvement in accuracy of the TOPMed imputation compared to 1000G. To illustrate how this reference panel can boost the discovery of complex disease variants we performed a type 2 diabetes GWAS meta-analysis using the TOPMed imputed data. TOPMed imputation not only improved the statistical significance of our findings but allowed for the testing of up to 24M rare variants, compared to 3M properly imputed with the 1000G panel. The high-quality of TOPMed imputation at low frequency and rare spectrum is especially relevant for the study of disease-causing variation, because deleterious variants usually span such frequencies. We show that by imputing with TOPMed, it is possible to test >90% of the variants with a MAF≥0.1% predicted to be deleterious by CADD score; these variants were previously only possible to detect by relying on more expensive sequencing technologies. While ascertaining variants at frequencies lower than 0.1% may still require WGS or WES, we estimate that the power to identify associated variants may be limited unless we undertake sequencing efforts with sample sizes orders of magnitude larger than our study. For example, for MAF<0.1%, the effective sample size required to reach sufficient statistical power to detect associations with an effect of OR>2.0 is above 170,000 individuals. Since the cost for sequencing such a large sample size is a major constraint for the study of underrepresented populations, we propose that highly accurate imputation with dense reference panels may be a more cost-effective approach.

In this study, we identified a novel low-frequency variant associated with type 2 diabetes, rs2891691, which lies between the *ORC5* and *LHFPL3* genes and showed increased accuracy of imputation and association power when using the TOPMed panel. *ORC5* encodes the subunit 5 of the origin recognition complex implicated in the DNA replication origins, and in transcription silencing and heterochromatin formation.[52] LHFPL3 is a member of the tetraspanin superfamily, which functions as membrane protein organizer. The rs2891691 risk allele is present in 1% frequency in Latino people. Carriers are associated with two-fold increased risk of developing type 2 diabetes, with a possibly higher risk in women.

By using the TOPMed panel, we identified a second low-frequency variant, rs1016378028, associated with a 1.7-fold increased risk of type 2 diabetes, which is not imputed with the 1000G panel. This variant was prioritized from a subset of variants at a sub genome-wide significant threshold that showed additional evidence of association. rs1016378028 is a Latin American private variant (MAF=1.3% in Latin America, MAF=0.2% in East Asia, MAF<0.05% in other populations), and lies within *HDAC2*, a gene highly intolerant of protein-changing variation that is mostly expressed in pancreatic islets.[33]

*HDAC2* is a member of the histone deacetylase family which catalyzes the removal of acetyl groups from histones, a chromatin modification involved in the repression of gene transcription. HDACs play a regulatory role in insulin signaling, β cell function and pancreatic endocrine cell development. For instance, at low glucose levels, HDAC2 is recruited to the insulin promoter to downregulate its expression.[53] Consistently, in human islets, the *HDAC2* expression negatively correlates with insulin gene expression (r=-0.56, FDR=3.7×10^−16^) and positively correlates with *IAPP* expression, which encodes for a hormone secreted after food intake to promote satiation (r=0.38, FDR=1.8×10^−7^).[34] HDAC2 also binds and deacetylates IRS-1 uncoupling its downstream phosphorylation cascade. Both insulin expression and signaling are partially restored after treatment with HDAC2 inhibitors.[54, 55] We show that cytokine treatment of pancreatic islets downregulated *HDAC2* expression. This may be a protective response, given that HDAC inhibition favors β cell development, shows anti-inflammatory effects and β cell protection against cytokine-induced apoptosis.[54, 55]

Because there is no comprehensive phenome-wide association data to guide the interpretation of variants enriched in Latin American populations, we assembled a QTL resource focused on glycemic and cardiometabolic traits by aggregating data from a total of 26,400 Latino individuals. This approach allowed us to follow up the identified signals and will be a valuable resource for future associations studies of Latino enriched variants with cardiometabolic traits. We observed that carriers of the rs1016378028 risk allele have higher levels of the ketone bodies, acetone 3-hydroxybutyrate and acetoacetate, which are produced through the breakdown of fatty acids and serve as an alternative energy source to glucose. Uncoupled hepatic production of ketone bodies may be a pathological consequence of relative insulin deficiency in diabetes.[56] 3-hydroxybutyrate also participates in the lysine β-hydroxybutyrylation, a histone mark that promotes transcription of starvation-responsive genes.[57] While the mechanism linking rs1016378028, diabetes risk and 3-hydroxybutyrate levels remains to be determined, our results suggest the rs1016378028 variant and *HDAC2* as a potential genetic type 2 diabetes risk factor.

We leveraged our GWAS results and existing publicly available data to develop an improved PS for Latino ancestry. PSs developed in a particular ancestry group poorly transfer to other populations, exacerbating disparities between populations. PS performance is greatly impacted by the accuracy of the effect sizes for each of the variants included in the GWAS summary statistics, which are influenced by the study sample size. We provide an improved PS model for Latino populations, by using a combination of GWAS and LD data from East Asian, European and our Latino GWAS. This PS showed a performance similar to the previously reported in European ancestry[58] with individuals at the top 2.5 percentile showing a four-fold increased risk of type 2 diabetes. Evaluating this PS in additional external datasets of Latino ancestry may prove useful in assessing its potential clinical utility.

Leveraging new resources to reanalyze Latin American data, such as imputation with the TOPMed reference panel, proved to be successful in identifying additional type 2 diabetes-related loci. Further efforts are needed to increase the power of discovery and to follow up on novel findings in diverse populations. Until then, translation of identified genetic variation-to-function and application to the clinic in Latin American populations will remain highly compromised compared to the resources available for European populations. For example, the lack of multi-omic data in publicly available biobanks including metabolically relevant tissues from individuals of Latin American descent, limits the characterization of the Latino-enriched or private novel signals. While in this study we gathered a high number of Latino samples with extensive biomarker and clinical characterization, larger sample sizes are still needed to achieve sufficient statistical power to detect low-frequency variants. Efforts must be expanded to build shareable resources with a high representation of different ancestries. This would enable ancestry-specific effects to be interpreted within the local ancestry context, which is instrumental to identify causal genes, to improve the biological mechanistic insight and to develop targeted therapies.

Overall, this study confirms the superior imputation performance of TOPMed, representing a cost-effective and unique opportunity to analyze low-frequency and rare variation in Latino samples at scale. It also presents the largest type 2 diabetes GWAS meta-analysis performed in individuals of Latin American ancestry imputed with the TOPMed reference panel. Despite the sample size being orders of magnitude smaller compared to studies performed in other populations, the novel discoveries presented here suggest that more novel genetic associations and new biology of type 2 diabetes will be found as the sample size of discovery samples, reference panels, and large-scale biobanks with phenome-wide data increase in studies including non-European populations.

## Data Availability

Full summary statistics will be made available through the Common Metabolic Diseases Knowledge Portal (https://cmdkp.org/), and the GWAS catalog. PRS weights for each ancestry will be shared via the PGS catalog (https://www.pgscatalog.org) and the Common Metabolic Diseases Knowledge Portal.

## Abbreviations

1000G: 1000 Genomes
CCHC: Cameron County Hispanic Cohort
em: expectation maximization
GERA: Genetic Epidemiology Research on Aging Cohort
LD: Linkage Disequilibrium
LoF: Loss-of-Function
METS: Mexican Metabolic Syndrome Cohort
MGB: Mass General Brigham Biobank
MXBB: Mexican Biobank Cohort
PAGE: Population Architecture using Genomics and Epidemiology
PRODIGY: Progress in Diabetes Genetics in Youth
PS: Polygenic Score
QTL: Quantitative Trait Locus
SIGMA: Slim Initiative for Genomic Medicine in the Americas Cohorts
T2D-GENES: Type 2 Diabetes Genetics Exploration by Next-generation sequencing in multi-Ethnic Samples
TODAY: Treatment Options for Type 2 Diabetes in Adolescents and Youth
TOPMed: NHLBI Trans-Omics for Precision Medicine
UKBB: UK Biobank
WES: Whole Exome sequencing

## AUTHORS’ RELATIONSHIPS AND ACTIVITIES

As of April 2022, P.D. is an employee and stockholder at Regeneron Pharmaceuticals. The rest of the authors declare no competing interests.

## CONTRIBUTION STATEMENT

A.H.-C. and J.M.M. conceived and planned the main analyses. A.H.-C., J.M.M., P.S., R.M., X.Y., M.C., J.C., A.L., J.D. and J.C.F. wrote and edited the manuscript. A.H.-C. and J.M.M. designed and performed the quality control. A.H.-C. performed the main analysis. M.U. and P.D. contributed with SIGMA3 phenotype harmonization. B.P. contributed with MGB phenotype harmonization. W.Z., L.P., J.McC., S.F., and J.B. contributed with CCHC data and analysis. R.G., L.L., R.S., M.K., and B.B. contributed with TODAY data and analysis. D.D., D.J., and S.M. contributed with SEARCH data and analysis. L.C., S.S., J.T., and J.F. contributed with T2DGenes data and analysis. R.L., B.D., C.K., L.R., C.H., Q.S., and H.H. contributed with PAGE data and analysis. R.H. contributed with Pima Indians data and analysis. P.S., A.M., and J.C. contributed with UK Biobank data and analysis. R.M., L.S., J.L., and A.M. contributed with AllofUs data and analysis. X.Y., D.L., P.M., L.M., and M.C. performed the human islets functional characterization. A.H.-C., A.D., R.M., J.J., K.L., and M.N. performed the PS analyses. D.D. and J.D. contributed with TOPMed linkage disequilibrium data. T.T., and C.A.-S. contributed with METS and MHTG data. T.T. and A.M. contributed with MXBB data. J.M.M., A.L. and J.C.F. supervised the study. All authors reviewed and approved the final manuscript.

