## Supplemental information for "The power of TOPMed imputation for the discovery of Latino enriched rare variants associated with type 2 diabetes"

### ELECTRONIC SUPPLEMENTAL MATERIAL (ESM)

### ESM METHODS

#### Discovery sample

We aggregated data from 6 Latino cohorts with a total sample size of 18,885 individuals (8,150 cases and 10,735 controls): SIGMA1, SIGMA2, SIGMA3, MXBB, MGB and GERA cohorts (Figure 1, Table S1).

The Slim Initiative for Genomic Medicine in the Americas (SIGMA) Cohorts [1–3] include individuals of Mexican or Mexican American origin recruited from both hospital- and population-based studies. The SIGMA study comprised of three different subsets. The SIGMA1 subset consists of 4,190 individuals, the SIGMA2 subset is composed of 3,730 individuals with a high proportion of Central and South America Native ancestry and the SIGMA3 subset includes 5,790 individuals. All human research was approved by the Comité de Ética e Investigación del Centro de Estudios en Diabetes, the Ethics and Research Committees of the Instituto Nacional de Ciencias Médicas y Nutrición Salvador Zubirán, the Ethics and Research Committes of the Instituto Nacional de Medicina Genómica and the Federal Commission for the Protection against Health Risks (COFEPRIS) (CAS/OR/CMN/113300410D0027-0577/2012).

The Mexican Biobank Cohort (MXBB)[4] includes population-based individuals from the 2000 Mexican National Health Survey conducted between the years 1999 and 2000 in Mexico. In the present study, we used a subset of 1,730 individuals for whom DNA genotyping data and type 2 diabetes definition were available. MXBB studies involving human participants were reviewed and approved by the Research Ethics Committee (Approval CI-1479) of the National Institute of Public Health, Mexico). The patients/ participants provided their written informed consent to participate in this study.

The Mass General Brigham (MGB) Biobank[5] maintains blood and DNA samples of patients seen at MGB hospitals, including the Massachusetts General Hospital, Brigham and Women's Hospital, McLean Hospital and Spaulding Rehabilitation Hospital, all in the Boston area (U.S.). We analyzed a subset of 2,115 genetically identified Latino samples. MGB work was conducted with approval from the MGB Institutional Review Board (study 2016P001018). It acknowledges the Partners HealthCare System for support of the MGB biobank and MGB patients for providing samples, genomic data, and health information data.

The Genetic Epidemiology Research on Aging cohort (GERA)[6] includes over 100K adults who were members of the Kaiser Permanente Medical Care Plan in the Northern California Region of the U.S. In this study, we analyzed a subset of 1,330 genetically identified Latino samples.

For each cohort, we selected Latino samples based on their genetically estimated ancestry, as described below. We calculated the principal components (PCs) using common genetic variants (minor allele frequency [MAF]≥5%) using FlashPCA[7] on both self-reported Latino individuals and the 1000G Phase 3 samples. Then, we separately estimated the mean of the top 4 PCs on either Latino samples or each of the four continental superpopulations defined in the 1000G Project (*i.e.* European, African, East Asian, and South Asian). For MGB and GERA, we applied a first filtering step to remove Latino samples within 2 SD of the mean for PCs 1-4 for each superpopulation cluster. This step was not applied to SIGMA and MXBB cohorts since they showed more homogeneous genetic ancestries that fall on the expected cline of Native American and European ancestry. For all discovery cohorts, we applied a second step where outlier Latino samples lying more than 4 SD away from the mean PC for its own Latino cluster were also removed. For all cohorts, we estimated the genetic-based global ancestry using Admixture[8] assuming five ancestral populations (k=5) (Figure S1).

**Genotyping, quality control and imputation**

Genotyping was done using several commercially available genome-wide arrays. A subset of the SIGMA samples (N=9,520) underwent whole-exome sequencing (WES), which we integrated with the genotyping array data to improve the imputation backbone (Table S1). We applied pre-imputation quality control to each dataset separately. Variant quality checks included the exclusion of variants with 5% or more missing data, MAF<0.1%, a Hardy-Weinberg equilibrium *P*-value$<5\times10^{-10}$, palindromic single nucleotide variants (AT or CG), duplicates, as well as a case *vs.* control missingness difference *P*-value<0.00005. We also excluded samples with 2% or more missing data, that did not pass the sex check or that were closely related as estimated by the proportion of identity by descent (IBD>0.185) between pairs of individuals. Clean datasets were phased using SHAPEIT2[9] and used as input for imputation.

For comparison purposes, we imputed the phased haplotypes using both 1000G Phase 3 version 5[10] and TOPMed reference panels freeze 8[11] in each cohort, separately. We used Michigan imputation and TOPMed imputation servers to impute genotypes.[12–14] For chromosome X, we imputed non-pseudoautosomal regions from females and males separately.

**Imputation performance evaluation**

We used the $r^2$ quality measure, as reported by the Michigan and TOPMed imputation servers. It calculates the ratio of the empirically observed variance of the allele dosage to the expected binomial variance.[15] We evaluated the performance of TOPMed and 1000G imputations by summarizing the chromosome-wise $r^2$ quality measure and the number of well-imputed ($r^2 \geq 0.8$) variants at different allele frequency thresholds. To further test if the quality of imputation was well-powered to detect low-frequency and rare variation without relying on the imputation server's quality measures, we leveraged available WES data from SIGMA3 cohort and estimated the proportion of the sequenced variants, for chromosome 22 only, that were well-imputed with TOPMed and 1000G panels at different WES allele frequency thresholds. Loss-of-Function (LoF) variants are of clinical interest as they disrupt protein-coding genes, potentially being disease-causal. Therefore, we used SnpEff[16] to annotate the WES variants for chromosome 22 only and estimated the percentage of well-imputed variants identified with TOPMed and 1000G imputations. We used CADD score[17] to predict the deleteriousness of variants. We classified the variants using the score cut-offs of 10 and 20, below which the variants were considered as benign.

We also calculated the effective sample size (N) needed to reach 80% statistical power to detect genome-wide significant associated signals ($\propto = 5\times10^{-8}$) at different effect sizes and allele frequencies covered by the imputations. Effective N was calculated as:

$$effective\ N = 4({}^{1}/_{N\ cases} + {}^{1}/_{N\ controls})$$

**Type 2 diabetes association and meta-analysis**

Type 2 diabetes association analyses were performed in each cohort with SNPTEST using the expectation maximization (*em*) method[18] for doing maximum likelihood estimation in a generalized linear model framework using genotype probabilities under the additive model. Models were adjusted for sex, age, body mass index (BMI) and 10 PCs to account for population structure. We ran additional models without adjusting for BMI. Only well-imputed variants ($r^2 \geq 0.5$) were meta-analyzed using the inverse of the corresponding squared standard errors in METAL.[19] The statistical significance threshold was set to $P<5\times10^{-8}$.

To extract the distinct type 2 diabetes associated signals, we clumped all variants with an association $P$-value$<5\times10^{-6}$. We set an LD $r^2=0.5$ and a distance between variants of 250 kb. If the variant was located within a previously reported type 2 diabetes-related *locus*, we used a conditioning strategy to test for distinct signals. We conditioned on each of the previously reported distinct SNPs within the *locus*. We performed conditional analyses in each cohort separately, and then meta-analyzed using the inverse of the variance of the effect estimates. If our lead SNP showed evidence of residual type 2 diabetes association ($P$-value$<5\times10^{-5}$) after conditioning on any previously reported SNPs within the *locus*, we considered our novel signal as distinct.

Distinct variants with sub-genome-wide significance ($P<1\times10^{-6}$) that were only imputed with the TOPMed reference panel, showed increased frequency in the Latino population and were > 250 kb from other reported genome-wide significant variants from large consortia analyzing either European or East Asian populations[20, 21] were considered for further investigation.

**Replication sample**

Variants associated with type 2 diabetes risk at genome-wide and sub genome-wide significance were tested for replication in six independent cohorts described below (Table S2).

*Progress in Diabetes Genetics in Youth (PRODIGY)*

PRODIGY is the largest cohort for youth-onset type 2 diabetes with available GWAS data, comprising the Treatment Options for Type 2 Diabetes in Adolescents and Youth (TODAY)[22] and the SEARCH for Diabetes in Youth studies[23]. TODAY includes participants diagnosed with type 2 diabetes prior to 18 years of age and have documented BMI≥85$^{th}$ percentile at the time of diagnosis. Of note, the American Indian tribal nations that partnered on the TODAY Study elected not to participate in the genomics collection. SEARCH is a population-based prospective registry study launched in 2000 that ascertained diabetes in youth diagnosed at <20 years of age in the U.S. Overall, PRODIGY has shown consistent direction and size of effects for most genetic variants associated with type 2 diabetes in adults.[24] The TODAY and SEARCH protocols were approved by the institutional review boards of each participating institution. Participants provided written informed parental consent and child assent, including consent and assent specifically for genetic testing.

We identified 1,198 youth type 2 diabetes cases of Latino descent. As the control group, we used 1,805 diabetes-free adult Latino samples from the T2D-GENES (Type 2 Diabetes Genetics Exploration by Next-generation sequencing in multi-Ethnic Samples) and from the METS (Mexican Metabolic Syndrome) Cohort.[25] Genotypes from both datasets were merged and the pre-imputation quality control additionally included the exclusion of variants with genotyping array missingness difference (*P*-value<0.00005). Phasing was done as described above and genotypes were imputed to the TOPMed reference panel to ensure high-quality imputation. Type 2 diabetes association was tested under an additive model using SNPTEST and *em* method[18]. Models were adjusted for sex and 10 PCs to account for population structure. Given the case-control design of this replication cohort, age and BMI were not considered as covariates.

*Cameron County Hispanic Cohort (CCHC)*

CCHC is a population-based cohort of Mexican American individuals living in the U.S.[26] We selected 971 type 2 diabetes cases and 857 controls. We used high-quality TOPMed imputed genotypes ($r^2$>0.9) and tested for type 2 diabetes association under additive models adjusted for sex, age, BMI and 10 PCs. Human research was approved by the relevant Institutional Review Boards. All participants provided written informed consent. CCHC was approved by the Committee for the Protection of Human Subjects at the University of Texas Health Sciences Center at Houston, Human Research Protections Program at Vanderbilt University.

*Urban American Indians and Arizona Pima Indians cohorts*

We also used 851 type 2 diabetes cases and 2,191 controls from four groups of urban-dwelling American Indians living in or near Phoenix, Arizona, as well as 2,571 type 2 diabetes cases and 5,088 controls from a community of Pima Indians in Arizona, who participated in a longitudinal study of type 2 diabetes.[27] For both cohorts, imputation was done using Pima whole-genome sequences from 266 individuals. Variant rs1016378028, was directly genotyped using Taqman probes. type 2 diabetes association was tested by fitting additive models adjusted for sex, age, BMI and 5 PCs. In the Pima cohort, we additionally adjusted for birth year since exams took place over many years. In the Pima cohort, we used linear mixed models to account for estimated relatedness among individuals, whereas in the Urban American Indians cohort, we used a genomic control procedure. Human research was approved by the relevant Institutional Review Boards. All participants provided written informed consent.

*Population Architecture using Genomics and Epidemiology (PAGE) study*

The PAGE study aims to conduct genetic epidemiological research in ancestrally diverse populations within the U.S.[28] It includes four population-based cohorts with significant numbers of Latino participants: the Hispanic-Community Health Study/Study of Latinos (HCHS/SOL), the Women's Health Initiative (WHI), the Multiethnic Cohort (MEC) and the Icahn School of Medicine at Mount Sinai BioMe biobank in New York City (BioMe). Genotyped individuals self-identified as Hispanic/Latino, African American, Asian, Native Hawaiian, Native American or other. For this study, Latino samples were selected based on their genetically estimated ancestry following the same two-step filtering approach that we implemented with the discovery cohorts. We analyzed up to 6,761 type 2 diabetes cases and 5,747 controls. We used high-quality TOPMed imputed genotypes ($r^2$>0.9) and tested for type 2 diabetes association under additive models adjusted for sex, age, BMI and 14 PCs. Analyses were conducted in SUGEN[29] to account for relatedness within datasets. Human research was approved by the relevant Institutional Review Boards. All participants provided written informed consent.

*All of Us Research Program*

The All of Us Program aims to build a national longitudinal resource of multiple data types and biosamples from at least one million individuals in the U.S., with the main goal of broadly reflecting the diversity in the country.[30] We analyzed whole genome sequencing data from 8,958 genetically identified Admixed American/Latino individuals, of which 1,243 were type 2 diabetes cases and 7,715 were controls. We tested the type 2 diabetes association under additive models adjusted for sex, age, BMI, and 16 PCs. All participants consent to participate. The work described here was confirmed as meeting criteria for non-human subject research by the *AoU* Institutional Review Board. All methods were carried out in accordance with relevant guidelines and regulations.

**Type 2 diabetes-related Quantitative Trait Locus (QTL) analyses**

Given the lack of large-scale publicly available biobanks with Latino samples that may allow for better characterization of our novel signals, including those occurring at a low frequency, we assembled a collection of cohorts to perform QTL analyses focused on 46 glycemic, anthropometric and lipid traits. In addition to 5 of the Latino cohorts analyzed in the type 2 diabetes meta-analysis (*i.e.* SIGMA1, SIGMA2, SIGMA3, MXBB and MGB Biobank), we included three extra cohorts, which we also imputed to the TOPMed panel as described above: the METS Cohort, the Mexican Hypertriglyceridemia (MHTG) Cohort, as well as the genetically identified Latino samples from the UK Biobank (UKBB).[31] The MHTG study was reviewed and approved by the Ethics and Research Committees from the Instituto Nacional de Ciencias Medicas y Nutricion Salvador Zubiran and UCLA Institutional Review Board MIRB1. The UK Biobank has obtained ethical approval covering this study from the National Research Ethics Committee (REC reference 11/NW/0382).

We also analyzed the Nightingale Nuclear Magnetic Resonance-based panel of 168 metabolomic biomarkers in Latino samples from the UKBB. The panel provides measures spanning multiple pathways, including lipoprotein lipids, fatty acids, amino acids, ketone bodies and glycolysis metabolites.

QTL analyses were conducted using high-quality TOPMed imputed genotypes and cleaned phenotypes from non-pregnant Latino adults. We used inverse rank normal transformation when the normality assumption was not met. Association analyses were done with a maximum of 26,400 adult Latino individuals depending on the trait, of whom 19,459 were diabetes-free. We used SNPTEST and *em* method to run linear regressions assuming additive genetic models in each cohort, separately. Models were adjusted for sex, age, BMI, and 10 PCs. If the outcome was available in >1 cohort, we meta-analyzed the results using the fixed-effects inverse variance method.

**Credible sets**

For each novel variant, we identified the set of variants with 99% probability of containing the causal variant. We used a Bayesian method[32], considering variants in LD with the lead variant ($r^2$>0.1). We calculated LD using genetic data from 1,996 Hispanic/Latino samples from TOPMed Freeze 5b. For each region, an approximate Bayes factor (ABF) was calculated for each variant as follows:

$$ABF = \sqrt{1 - r\, e^{\left(\frac{rz^2}{2}\right)}}$$

where $r = {}^{0.04}/_{(SE^2 + 0.04)}$ and $z = {}^{\beta}/_{SE}$

The $\beta$ and the SE are the estimated effect size and the corresponding standard error, respectively, that result from testing the variant association under a logistic regression model.

The posterior probability for a variant being causal is equal to its ABF divided by the sum of all ABF values for the *locus*. Large values of the Bayes factor correspond to strong evidence for association. Therefore, variants are ranked by ABF in decreasing order, and the cumulative probability is calculated starting at the top of the list until the value exceeds 99%.

**Genomic annotation of the 99% credible set variants**

We used the 99% credible sets for each novel signal to annotate their genomic effect using the VEP[33] (GRCh38.p7) and SNPNEXUS[34] applications. This allowed us to gather information about gene and protein proximity, the effect of non-synonymous coding variants on protein function (SIFT, PolyPhen), non-coding variants scoring (CADD, FunSeq2), and the occurrence of regulatory elements.

We used GTEx V8[35] to assess the influence of the variants in gene-level expression, as well as the TIGER Portal[36] for evaluating the gene-level expression in pancreatic islets and the Islet Gene View[37] for assessing the gene co-expression in human islets. We also assessed individual variant associations with a variety of common phenotypes and diseases using the Common Metabolic Disease Knowledge Portal (cmdgenkp.org. 2021 Nov 15), as well as other resources, including web servers such as UK Biobank imputed with TOPMed (https://pheweb.org/UKB-TOPMed/) and FinnGen (https://r6.finngen.fi/). We also assessed gene-phenotype associations using the Genebass browser[38], a resource of exome-based association statistics across 281,852 individuals with exome sequence data from the UKBB.[39]

To obtain more evidence implicating the variants or their closest genes, with any disease or biological process, we used the Open Targets Platform, which aggregates a variety of resources and scores the collected information to contextualize and weigh the underlying evidence.[40]

**Expression of genes near novel variants**

We also assessed the expression levels of the genes ± 500 kb around the novel signals in human islets under different conditions pertaining to type 1 diabetes and type 2 diabetes. We examined the following datasets: Type 1 diabetes dataset (FACS-purified β cells obtained from donors with type 1 diabetes) downloaded from GSE121863 (n=4 type 1 diabetes versus 12 controls)[41]; type 2 diabetes samples gathered and integrated from three cohorts: one generated by T2DSystems (TIGER Portal), and the other two downloaded from the Gene Expression Omnibus (GEO) database under accession numbers GSE159984 and GSE50244 (a total of n= 47 type 2 diabetes cases versus 228 controls)[36, 42, 43]; brefeldin A-exposed human islets (0.1 μg/mL for 24h) downloaded from GEO under accession number GSE152615 (n=4)[44]; cytokine-exposed human islets (exposed to interferon-γ (1,000 U/ml) and interleukin-1β (50 U/ml) for 48 h or to interferon-α (2000 U/mL) for 2h, 8h, or 18h, n=5-6 per condition) from GSE108413 and GSE133221[45, 46]; human islets exposed to palmitate (0.5 mM), high glucose (22.2 mM) and palmitate plus high glucose for 48h (n=3-5 per condition) from GSE159984[42]. Gene expression differences between groups were assessed using *P*-values and adjusted *P*-values (Benjamini Hochberg correction) determined by the Wald test using the DESeq2 pipeline.[47] Transcript per million (TPM) was normalized by Salmon 1.4.0.[48]

**Polygenic scores**

Polygenic scoring using single ancestry summary statistics and LD reference panels was calculated via Bayesian Regression and Continuous Shrinkage priors as implemented in PRS-CS.[49] We used the UKBB LD reference panel and GWAS summary statistics from European[20], East Asian[21] and Latin American populations. GWAS Latino summary statistics were calculated using a meta-analysis with five of the discovery cohorts (*i.e.* SIGMA1, SIGMA2, SIGMA3, MGB, and GERA). Five phi shrinkage priors ($\Phi$) were used (*i.e.* 1, $10^{-2}$, $10^{-4}$, $10^{-6}$ and the one automatically estimated from the data). Then, we used the estimated posterior SNP effect sizes for each ancestry to calculate the PSs in a training cohort (*i.e.* MXBB).

To evaluate the performance of the PSs, we first fitted a simple model that included sex, age and 10 PCs to account for population stratification. We then fitted models that additionally included the PS standardized scores. We calculated the variance explained in type 2 diabetes status for each model using the Nagelkerke pseudo $R^2$. We repeated the same strategy by comparing the individuals above different percentile cutoffs with those in the interquartile of the PS. The best shrinkage prior was selected based on the larger incremental $R^2$ of type 2 diabetes status in the MXBB training cohort. Then, the selected model was tested in a target cohort (*i.e.* the METS Cohort).

Given that the ancestry-specific PSs were not highly correlated ($r^2$<0.3), we also used PRS-CSx[50], a novel method that improves cross-population polygenic prediction by integrating GWAS summary statistics from multiple populations. For a given shrinkage prior, PRS-CSx returns posterior SNP effect size estimates for each discovery population, which we used to calculate the cross-population PS. First, we fitted a linear regression combining the standardized scores for each population in the MXBB training cohort, as follows:

$$y \sim B_{\emptyset,Pop1}PRS_{\emptyset,Pop1} + B_{\emptyset,Pop2}PRS_{\emptyset,Pop2} + \cdots + B_{\emptyset,K}PRS_{\emptyset,K}$$

where y is type 2 diabetes status, $B_{\Phi,Pop}$ is the regression coefficient for a given phi shrinkage prior and population and $PS_{\Phi,Pop}$ is the standardized PS for a given phi shrinkage prior and population. The phi shrinkage prior and corresponding regression coefficients for the linear combination of PS that maximizes the incremental Nagelkerke pseudo $R^2$ were used in the METS target cohort to evaluate the performance of the cross-population PS.

$$PRSCSx = \hat{B}_{\emptyset,Pop1}PRS_{\emptyset,Pop1} + \hat{B}_{\emptyset,Pop2}PRS_{\emptyset,Pop2} + \cdots + \hat{B}_{\emptyset,K}PRS_{\emptyset,K}$$

We also calculated the AUC either for the covariates sex, age and 10 PCs or the cross-population PS plus the covariates. The DeLong test was used to assess the difference between AUCs. We then calculated the OR per standard deviation in the cross-population PS, adjusting for sex, age and 10 PCs. Finally, we identified the high-risk individuals at the top 2.5%, 5% and 10% of the cross-population PS distribution and calculated the OR of the high-risk individuals *versus* the interquartile distribution.

**ESM FIGURES**

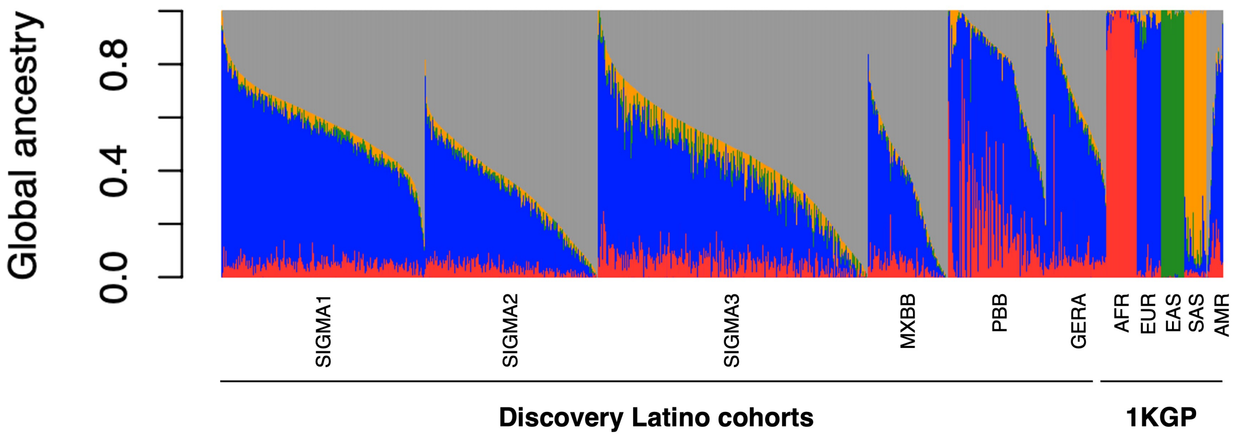


**ESM Fig. S1. Individual global ancestry proportions in the discovery Latino cohorts.** Global ancestry proportions were calculated after merging genotypes with 2,504 individuals from the 1000G phase 3, using ADMIXTURE software at K=4. Colors represent the super-continental ancestries: African (red), European (blue), East Asian (green) and South Asian (yellow). Grey color represents Admixed American ancestry.

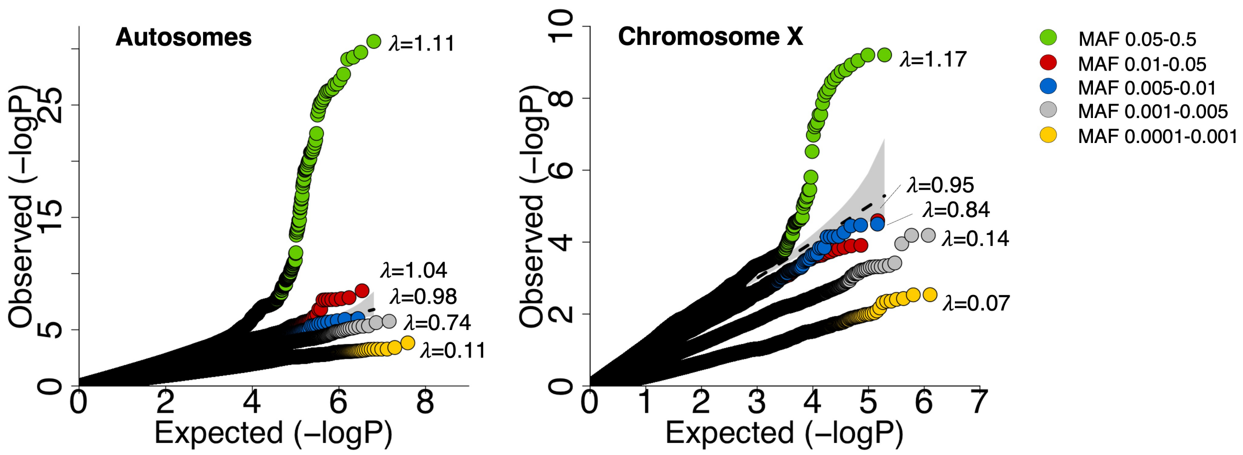


**ESM Fig. S2. QQ plots of association statistics in GWAS.** Plots show the calibration under the null and enrichment of T2D-associated variants in the tail for autosomes and chromosome X, stratifying by minor allele frequency. There was a minimal inflation of test statistics for variants with an allele frequency of 0.05 or higher, as indicated by a slight early departure between observed and expected *P*-values of the QQ plots (lambda=1.1). For low-frequency or rare variants, we observed deflated QQ plots, which is expected given the large sample sizes needed to reach statistical power.

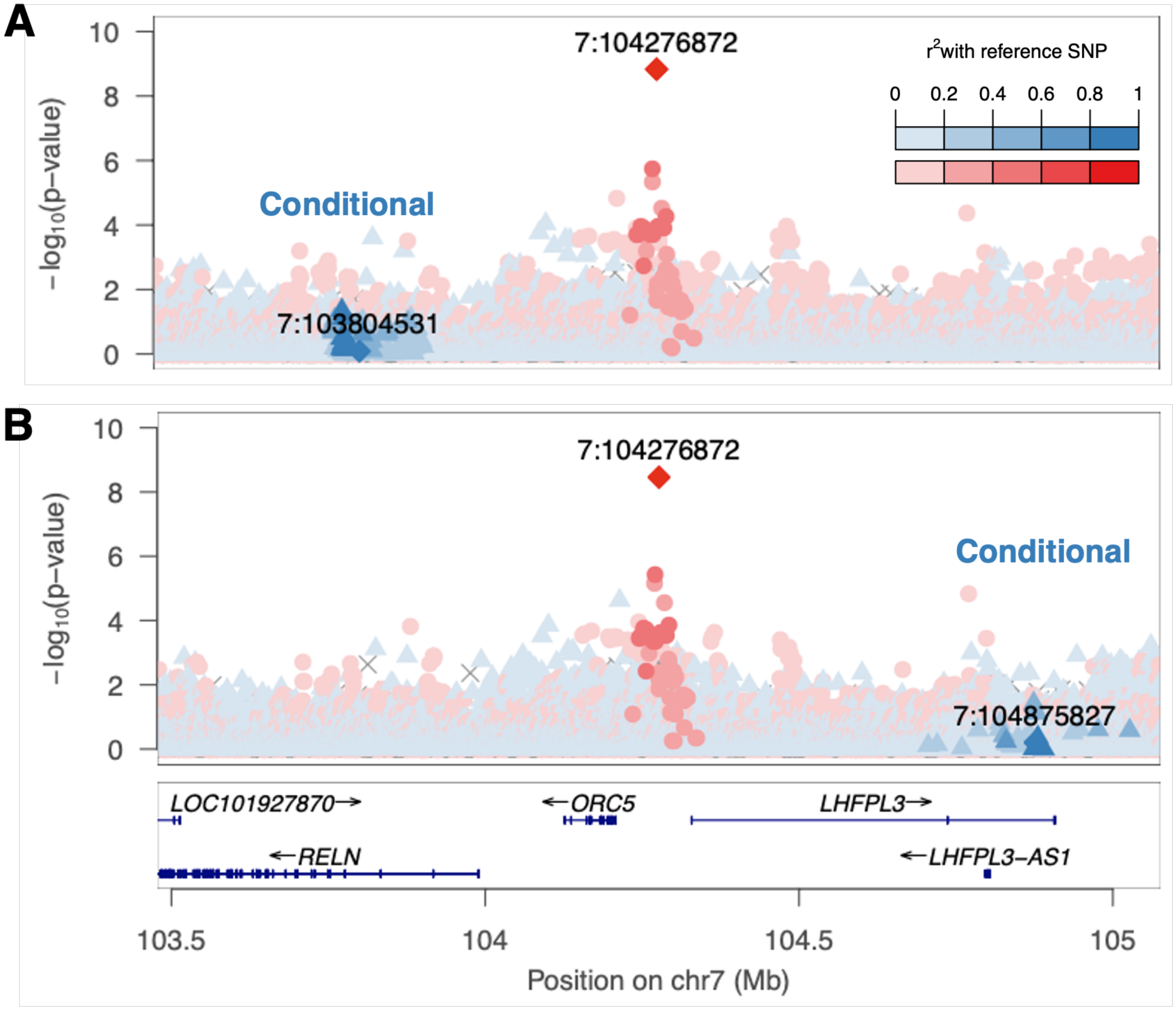


**ESM Fig. 3. Regional plot of the association at novel *ORC5/LHFPL3 locus* conditional on two nearby T2D-associated signals.** Two type 2 diabetes-associated signals near the rs2891691 variant have been reported in Europeans. One is located within the upstream *RELN* gene (rs39328, b38 chr7:103,804,531)[51], while the other is located within the downstream *LHFPL3* gene (rs73184014, b38 chr7:104,875,827)[52]. rs2891691 was not in LD with either of these variants ($r^2$<0.0006), and neither of them was associated in our Latino meta-analysis (*P*=0.30 and *P*=0.07, respectively). After conditioning on each of them, rs2891691 remained significant (OR [95% CI] =2.01 [1.59-

2.53], $P$=3.1×10$^{-9}$ and OR [95% CI] =2.01 [1.59-2.53], $P$=3.5×10$^{-9}$, respectively). Red color intensity indicates $r^2$ to the novel variant rs2891691. Blue color intensity indicates $r^2$ to the conditional variants.

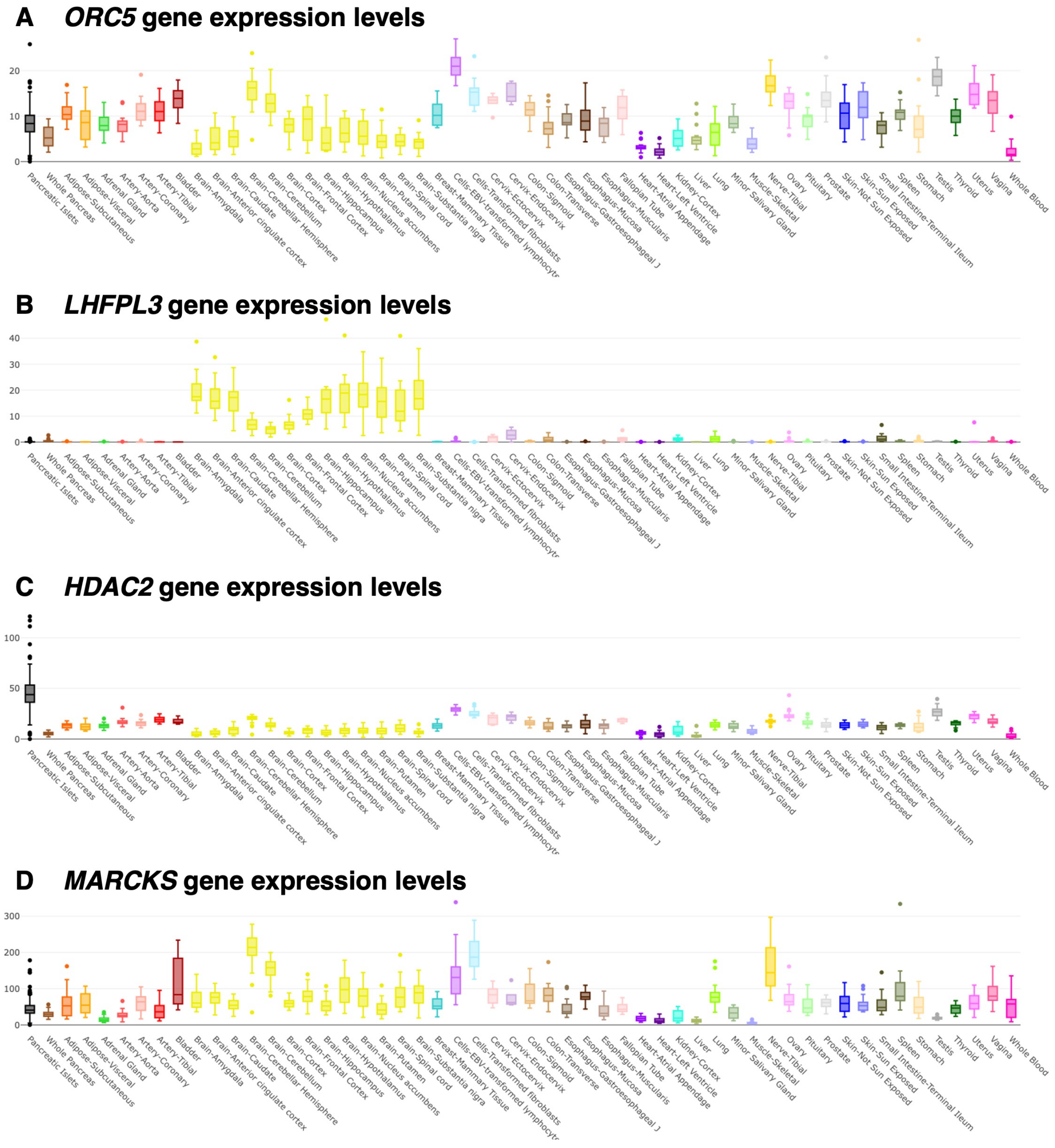

**ESM Fig. 4. Expression levels of genes around the two Latino T2D-associated leading variants.** Multiple tissue and human islets expression levels (TPM) from GTEx and TIGER portals.

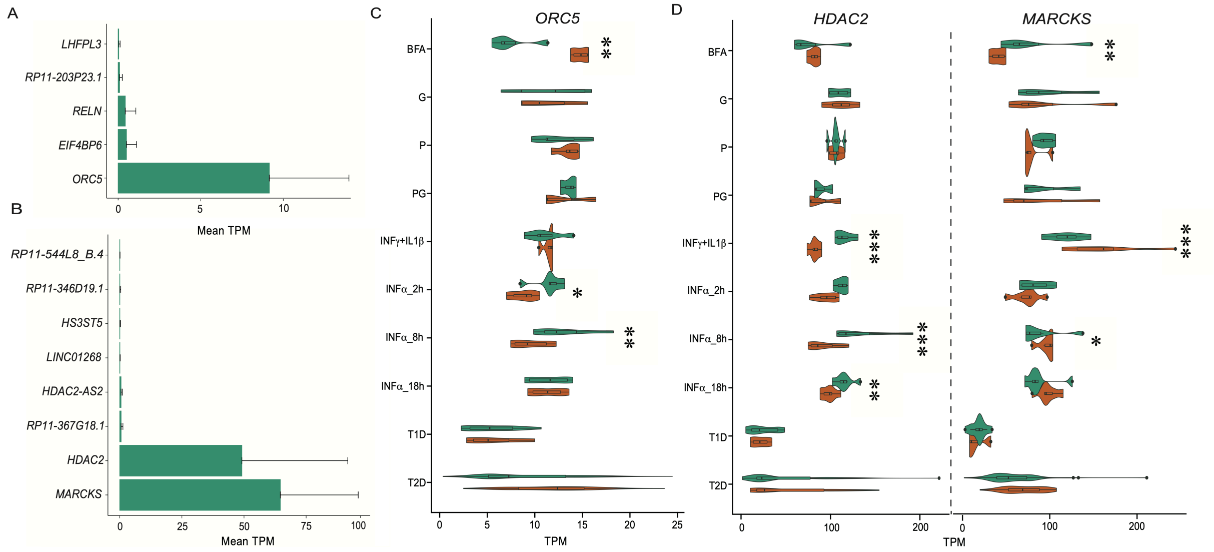


**ESM Fig. 5. Expression levels of genes around the two Latino T2D-associated leading variants in human islets under different conditions.** a. Expression levels (mean TPM+SD) of genes ± 500 Kb to *ORC5* lead variant in human islets under control condition. b. Expression levels (mean TPM+SD) of genes ± 500 Kb to *HDAC2* lead variant in human islets under control condition. c. Expression level of *ORC5* and d. *HDAC2* and *MARCKS* in human islets under different conditions (in orange, compared to control in green) and in islets of patients with T1D or T2D. Asterisks show the adjusted (Benjamini Hochberg correction) p value * <0.05; ** <0.01; *** <0.001. BFA brefeldin A, G high glucose, P palmitate, PG palmitate + high glucose; specific conditions and data sources are provided in the Methods section.

### ESM REFERENCES

health and aging (GERA) cohort. Genetics 200(4):1285–1295. https://doi.org/10.1534/genetics.115.178616

cohorts. Diabetologia 62(12):2298–2309. https://doi.org/10.1007/s00125-019-05001-w

human pancreatic beta cells. Nature Communications 2020 11:1 11(1):1–17. https://doi.org/10.1038/s41467-020-16327-0

multi-ancestry meta-analysis. Nat Genet 52(7):680–691. https://doi.org/10.1038/s41588-020-0637-y
